# Associations Between Air Pollution and relative Leukocyte Telomere Length Among Northern Swedish Adults: Insights from the Betula Study

**DOI:** 10.1101/2024.03.10.24304057

**Authors:** Wasif Raza, Sara Pudas, Katja M. Kanninen, Erin Flanagan, Sofie Degerman, Rolf Adolfsson, Rosalba Giugno, Jan Topinka, Xiao-wen Zeng, Anna Oudin

## Abstract

Air pollution is increasingly discussed as a risk factor for dementia, but the biological mechanisms are not yet fully understood. Cellular integrity markers like telomere length are relevant to investigate in relation to air pollution exposure in this context, as they are associated with aging and dementia. Previous studies on air pollution and telomere length have somewhat mixed results, which may partly be due to differences in air pollution modelling, composition, and concentrations. The study aimed to investigate the relationship between source-specific air pollution exposure and telomere length in a low-level air pollution area.

Data were obtained from the Betula project, a longitudinal study in Northern Sweden dedicated to studying aging, memory and dementia. A total of 519 participants who were recruited between 1988 and 1995 were included, whose relative telomere length was measured, and who were followed-up with respect to dementia for more than 20 years. To estimate annual mean air pollution concentrations, a dispersion model linked to each participant’s residential address was employed. We conducted linear regression to explore the associations between annual mean air pollution concentrations at participants’ residences and relative leukocyte telomere length.

In the whole sample, there was no support for air pollution to affect telomere length, with regression slope estimates close to zero and p-values >0.10. There were tendencies for a positive association of longer telomere length and higher exposure to air pollution among individuals who were later diagnosed with dementia (N = 74), but these findings were not conclusive (p-values >0.10). The findings raise questions about susceptibility to air pollution and the state of the inflammatory response in individuals who later will develop dementia. Given the imprecise nature of these estimates, caution is advised in their interpretation however, and validation in other studies is essential.

## 1. Introduction

Ambient (outdoor) air pollution is a major global health concern (1). The World Health Organization (WHO) estimates that ambient air pollution causes 3.5 million premature deaths annually and that 99% of the world’s population are currently being exposed to levels above the current air quality guidelines outlined by WHO in 2021 (2). Even at relatively low concentrations, adverse health effects of air pollution thus persist, highlighting its ongoing harm to human health (3).

Extensive evidence from large-scale studies and meta-analyses has demonstrated associations between exposure to ambient particulate air pollution, and the development of dementia (4). With an ageing population, dementia is now the leading cause of death in some European countries, for example the UK and Belgium. Alzheimer’s disease (AD) stands as the primary cause of dementia and is characterized by various pathological features, including the buildup of amyloid-β plaques and neurofibrillary tau tangles, which result from complex interactions among genetic and lifestyle factors (4). Given the absence of a definitive therapy for AD and other dementia disorders, it is imperative to identify biomarkers that can help predict an individual’s susceptibility to the disease. Notably, air pollution, as a modifiable environmental risk factor, presents an opportunity to address the risk of dementia (4, 5). It is thus furthermore imperative to understand the role of air pollution on such potential biomarkers.

With the ultimate goal of preventing and better treatments for AD and other dementia disorders, a deeper understanding of the molecular-level impact of air pollution and its effects on cellular biomarkers is crucial. Telomeres, the non-coding ends of chromosomes, play a pivotal role in maintaining genomic stability and integrity by safeguarding the coding genome (6). These protective caps prevent chromosome erosion and fusion, but successively shorten during DNA replication and cell division. Comprised of repeated DNA sequences and specialized proteins, telomeres form a complex structure crucial for cellular health and longevity (7). Accumulating evidence has linked shorter relative leukocyte telomere length (rLTL) with aging, AD (8), age-related diseases and mortality (9). Research indicates that telomeres undergo progressive shortening with age, but various environmental and lifestyle factors can expedite this process. The harmful effects of air pollutants are rooted in inflammation, characterized by oxidative stress and systemic inflammation, raising concerns about their potential impact on cellular integrity and aging as well as overall health. Unravelling the effects of air pollution on telomere length can thus provide insights into the underlying mechanisms through which environmental factors influence human health and disease progression.

According to a systematic review of 12,058 subjects from 2018, exposure to air pollution (long-term, short-term or occupational exposure) may shorten telomere length (10). In a systematic review from 2019, only four studies were deemed high enough quality for a meta-analysis however, and the meta-estimate was inconclusive (11).

Health effects of air pollution in general, and inflammatory effects specifically, can be expected to differ depending on the composition of air pollution, both in terms of dominating sources of local air pollutants and the proportion of pollution coming from near sources or from long distance transport. It has for example been suggested that air pollution from sources nearby is more harmful for health than regional-level air pollution (12). To date, no previous study on air pollution and telomere length has distinguished between different sources of air pollution, or between local or regional air pollution. Studies in areas with low levels of air pollution and high-resolution air pollution models are furthermore lacking.

This study aims to address this gap by exploring the association between air pollution and rLTL in a low-level air pollution area while distinghuishing between different sources of local air pollution, and between local and total air pollution (where the latter include air pollution from long-range transport).

## 2. Material and Methods

### 2.1. Study population

The Betula cohort is a longitudinal, population-based study on dementia, memory and ageing which was initiated in 1988 to investigate health and cognitive trajectories in a representative fraction of the adult and elderly population residing in Umeå county, located in the Västerbotten region of Sweden. The comprehensive recruitment process for the Betula study has been extensively detailed in previous publications (13). In summary, participants in the Betula cohort have undergone examinations up to seven different time waves (T1 to T7), spaced at five-year intervals from 1988 to 2017. Each assessment includes the administration of health-related questionnaires and examinations and cognitive evaluations. For the purposes of the present study, participants enrolled during the time periods of 1988 to 1990 (test wave 1; T1) and 1993 to 1995 (test wave 2; T2) were selected. i. In the Betula cohort, diagnoses of dementia was established adjacent to each test wave to discriminate incident dementia cases from healthy individuals or individuals with cognitive impairment of non-progressive nature and to define the year of disease onset. The diagnostic procedure has previously been extensively described (14, 15).

### 2.2. Relative leukocyte telomere length

The relative leukocyte telomere length (rLTL) was measured by quantitative polymerase chain reaction (qPCR) in DNA from whole blood from the second test wave (T2) between 1993 and 1995 for a total of 519 included participants. The method for rLTL measurement by qPCR was originally described by Cawthon in 2002 (16). In this study, this method was used with some minor modifications (17). Each sample was evaluated by Telomere (TEL) and single copy gene hemoglobin subunit beta (HBB, Gene ID:3043) PCR reactions. A TEL/HBB value were calculated using the 2^−ΔCt^ method, in which ΔCt = average Ct_TEL_-average Ct_HBB_. The rLTL value were obtained by dividing the TEL/HBB value of each sample with the TEL/HBB value of a reference cell line (CCRF-CEM) DNA included in all runs. The rLTL values were further normalized for potential plate effects (between-run variability) by subtracting mean-centered plate effects, estimated through a mixed-effects model with age, gender, age × gender interaction and plate as fixed effects, and individuals as a random effect.

### 2.3. Air pollution exposure assessment

The annual mean concentrations of fine particulate matter (PM2.5) and Black Carbon (BC) were computed by the Swedish Meteorological and Hydrological Institute (SMHI) (18). Local and regional emission inventories served as inputs for the national dispersion modelling system, and SMHI provided the necessary data for the Gaussian dispersion model simulations to calculate annual mean concentrations of PM2.5 and BC. The model exhibited high spatial resolution, with concentrations modelled near major roads and close to smokestacks down to 35 m by 35 m. Additionally, the model incorporated emissions from industrial sources and shipping as point sources in its simulations. Emission factors for various types of vehicles were determined based on the Handbook on Emission Factors for Road Traffic, version 3.1 (19). Non-exhaust emissions, which encompass road wear and some contributions from brake wear and tire emissions, were also quantified. To estimate emissions from residential wood combustion, data from inventories of individual stoves and boilers, along with information gathered from chimney sweepers and interviews regarding wood burning habits, were utilized (20). The assessment of long-range transport air pollution relied on data from rural background monitoring stations. This involved calculating the difference between the total concentrations measured at these monitoring stations and the modelled local particle concentrations at the same locations.

Finally, the resulting concentrations of air pollution were linked to each participant’s residential address at sampling, and the modelled value of year 1990 was used as marker for long-term exposure for the participants. The six exposure variables included total concentrations of PM2.5 and BC (PM2.5_total, BC_total) as well as source-specific concentrations of PM2.5 (PM2.5_exhaust, PM2.5_woodburning) and BC (BC_exhaust, BC_woodburning). The study area, Umeå municipality, is located in Northern Sweden, and has overall quite low levels of PM2.5, with a total level of 9.81 µg/m3 in the present study (Table 1). This is below the former WHO recommended air quality guideline from 2005 of 10 µg/m3, but over the revised WHO (in 2021) air quality guideline of 5 µg/m3.

**Table 1.** Characteristics of the participants.

| Variables | Number of observations | Values<sup>a</sup> |
| --- | --- | --- |
| Age | 473 | 61.7 (14.7) |
| Gender |  |  |
| Female | 251 | 53.1 |
| Male | 222 | 46.9 |
| Education level |  |  |
| Compulsory | 158 | 33.4 |
| High school | 223 | 17.4 |
| University | 90 | 27.0 |
| Smoking status |  |  |
| Former smoker or Smoker | 224 | 47.4 |
| Non-smoker | 247 | 52.2 |
| <sup>b</sup>Lymphocyte proportion | 461 | 0.30 (0.08) |
| <sup>c</sup>PM2.5_total | 473 | 9.81 (0.66) |
| <sup>d</sup>PM2.5_exhaust | 473 | 0.24 (0.25) |
| <sup>e</sup>PM2.5_wood | 473 | 1.24 (0.31) |
| <sup>f</sup>Black C_total | 473 | 0.56 (0.15) |
| <sup>g</sup>BC_exhaust | 473 | 0.11 (0.12) |
| <sup>h</sup>BC_wood | 473 | 0.16 (0.04) |
| Dementia status |  |  |
| Dementia | 75 | 15.9 |
| No dementia | 398 | 84.1 |

<sup>a</sup> Values are mean (standard deviation) for continuous variables, percentage for categorical variables. <sup>b</sup> Lymphocyte proportion was calculated as lymphocyte count divided by the sum of all white blood cells count (sum of neutrophils, eosinophils, basophils, lymphocytes, and monocytes); <sup>c</sup> Total PM2.5 (particulate matter with an aerodynamic diameter of $\leq 2.5$ micrometers, “fine” particulate matter) concentration; <sup>d</sup> Particle emission from vehicles on the road; <sup>e</sup> Particle emission from wood burning; <sup>f</sup> Total black carbon concentration; <sup>g</sup> Black carbon emission from vehicles on the road; <sup>h</sup> Black carbon emission from wood burning.

### 2.3. Statistical analysis

The calculation of blood lymphocyte proportion involved dividing the lymphocyte count by the total count of white blood cells, which encompasses neutrophils, eosinophils, basophils, lymphocytes, and monocytes. To mitigate the influence of age and gender-related variations, relative leukocyte telomere length (rLTL) was residualized against the participant’s age at the time of rLTL measurement and their gender, using a linear regression model. This yielded a residualized telomere length, denoted as rLTL, which was then utilized to investigate the relationship between air pollution and telomere length. We considered age, gender (female, male), lymphocyte proportion, education level (compulsory, high school, university), and smoking status (smoker or former smoker, non-smoker) as a potential confounding factors to be included in this study. Air pollution exposure, rLTL as well as all potential confounders were assessed at test wave T2, whereas dementia was assessed with follow-up to February 2022.

Linear regression analysis was employed to assess the impact of both total and source-specific particle concentrations on rLTL. Model 1 was an unadjusted model. Model 2 incorporated adjustments for lymphocyte proportion, age and gender. Model 3 (the main model) additionally accounted for individual-level baseline potential confounders, including education level and smoking status. Furthermore, subgroup analysis based on future dementia diagnosis (Dementia, No dementia) was carried out to evaluate whether the association between air pollution and rLTL differed depending on dementia status. Here, a linear regression model was used and a multiplicative interaction term was included in the main model between each of pollutants and dementia status to explore potential effect modification. All statistical analyses were performed using R version 3.4.8.

The study was approved by the ethics review authority with Dnr: 2022-04608-01 and written informed consent was given by all Betula participants. The data for the present study were accessed for research purposes on the 15^th^ of March 2023. The researchers analysing the data did not have access to information that could identify individual participants.

## 3. Results

In the Betula study sample, 519 individuals had their rLTL measured during T2. Furthermore, residualization of rLTL against age and gender varibles resulting in a refined dataset consisting of 473 participants for initial analysis. Detailed characteristics of these participants can be found in Table 1.

The age of the participants ranged from 40 to 85 years (mean: 61.7 years, standard deviation (SD): 14.7). Among this group, 53.1% were female, 47.4% were smokers, and 27% had completed a university degree. The annual mean concentrations (SD) of total PM2.5 and BC were 9.83 (0.64) μg/m^3^ and 0.562 (0.143) μg/m^3^, respectively for 1990 (used as marker for long-term exposure in the study). During the follow-up period up to 2016, 15.9% developed dementia up to 2022, and the proportion of females in the dementia group was higher (53%) than the proportion of males (47%). The mean rLTL value exhibited variation depending upon whether participants would later be diagnosed with dementia or not. Shorter mean rLTL values (−0.0072) were observed among study participants who later were diagnosed with dementia than those who were not (0.00135), however, the difference was not statistically significant (p-value= 0.61) The linear associations between air pollution and residualized leukocyte telomere length (rLTL), are presented in Table 2.

**Table 2.** Linear associations between pollution exposure and relative leukocyte telomere length, model coefficients (β) with their 95% Confidence Intervals (CIs).

| Measures | Model 1<br>(N=473) | Model 2<br>(N=462*/461**) | Model 3<br>(N=460*/459**) |
| --- | --- | --- | --- |
| Air pollution exposure ($\mu\text{g}/\text{m}^3$) | $\beta$ (95% CI) | $\beta$ (95% CI) | $\beta$ (95% CI) |
| PM2.5_total | 0.01 (-0.009, 0.026) | 0.01 (-0.011, 0.024) | 0.001 (-0.011, 0.025) |
| PM2.5_exhaust | 0.03 (-0.019, 0.074) | 0.03 (-0.023, 0.072) | 0.02 (-0.023, 0.072) |
| PM2.5_woodburning | 0.01 (-0.025, 0.049) | 0.004 (-0.33, 0.042) | 0.001 (-0.033, 0.044) |
| BC_total | 0.04 (-0.036, 0.119) | 0.04 (-0.042, 0.116) | 0.03 (-0.046, 0.114) |
| BC_exhaust | 0.06 (-0.039, 0.155) | 0.05 (-0.046, 0.152) | 0.05 (-0.019, 0.042) |
| BC_woodburning | 0.13 (-0.194, 0.452) | 0.11 (-0.165, 0.432) | 0.11 (-0.165, 0.428) |

Model 1: Unadjusted; Model 2: Model 1 + Age, gender and lymphocyte proportion; Model\_3: Model 2 + smoking and education. \* No of observation in models with PM2.5 exposure variable. \*\* No of observation in models with BC exposure variable.

Notably, small but imprecise tendencies for rLTL to increase with air pollution exposure were observed, especially for the BC elements. For example, higher concentrations of BC were associated with longer rLTL: a 1 μg/m³ increase in total BC was associated with a unit increase of 0.04 (95% CI:-0.046, 0.114) in rLTL in Model 3 (Table 2). The corresponding estimates for PM2.5_exhaust, PM2.5_woodburning, BC_traffic and BC_woodburning were: 0.02 (95% CI: −0.023, 0.072), 0.001 (95% CI: −0.033, 0.044), 0.05 (95% CI: −0.019, 0.042), and 0.11 (95% CI: −0.221, 0.445), respectively. The estimates from the subgroup analysis based on dementia status are reported in Table 3.

**Table 3.** Associations between air pollution and relative leukocyte telomere length stratified by dementia (Model 3, residualized model coefficients (β) and p-values for interaction between pollutants and dementia status).

|  | Dementia<br>(74) | No dementia<br>(385) |  |
| --- | --- | --- | --- |
| Air pollution exposure<br>($\mu\text{g}/\text{m}^3$) | $\beta$ (p-value)<sup>a</sup> | $\beta$ (p-value)<sup>a</sup> | p-value |
| PM2.5_total | 0.03 (0.26) | -0.002 (0.87) | 0.14 |
| PM2.5_exhaust | 0.06 (0.14) | 0.004 (0.89) | 0.24 |
| PM2.5_woodburning | 0.034 (0.48) | 0.0004 (0.99) | 0.47 |
| BC_total | 0.11 (0.17) | 0.003 (0.96) | 0.20 |
| BC_exhaust | 0.12 (0.13) | 0.001 (0.88) | 0.28 |
| BC_woodburning | 0.41 (0.35) | 0.073 (0.70) | 0.59 |

<sup>a</sup>Adjusted for age, gender, lymphocyte proportion, smoking status and education level.

Associations between air pollution exposure and rLTL appeared to be present mainly in study participants who later developed dementia, but all p-values are larger than 0.10, so the results are not conclusive. The interaction terms for effect modification by dementia status were furthermore not statistically significant.

## 4. Discussion

In this study, associations between air pollution and telomere length were not evident. There were however some tendencies for telomere length to increase with exposure to ambient particles, among study participants who were later diagnosed with dementia. Overall, the results diverge from our initial hypothesis however, which anticipated air pollution to be associated with a *decreased* telomere length. The statistical precision of the estimates were low. Consequently, these findings should be interpreted cautiously and corroborated in other studies. If we speculate however, the results raises questions about susceptibility to air pollution and about the state of the inflammatory response in people who develop dementia. The idea that susceptibility to air pollution may differ between different groups in the population is not new; we have for example previously observed that associations between particulate air pollution and dementia were mainly present among study participants carrying the *APOE* ɛ4 allele and for those with low performance on odor identification ability in the same cohort (21).

Previous research on air pollution and telomere length have yielded somewhat conflicting findings although long-term exposure (10, 11). In a study using cross-sectional data on 471,808 UK Biobank individuals, telomere length decreased with increases in long-term concentrations of air pollution (22). Another large study conducted on UK Biobank participants found no significant association between long-term exposure to several pollutants and telomere length however (23). Discrepancies may thus occur in the same cohort, the main difference between the two studies seem to be the air pollution modelling. In alignment with our findings, a study involving school children in East London demonstrated an increase in telomere length with rising long-term concentrations of air pollution (24). Interestingly, genetic ancestry seemed to influence the associations in that study, with stronger associations being observed in black children. In the present study, associations seemed stronger in people who later developed dementia, which is also linked to genotype. In a study from Shanghai, China, on patients with diabetes mellitus, no associations between short-term levels of air pollution and telomere length was observed (25). In a heterogenous cohort of critically ill patients it was observed that long-term exposure to air pollution was s positively associated with telomere length (26). The authors speculate that an interaction of air pollutant exposure and acute inflammation may activate telomerase in these patients (26).

The mixed findings in previous studies underscore the complexity of the relationship between air pollution and telomere length, suggesting that multiple factors may contribute to the observed results. The levels and composition of air pollution may evidently partly explain some of the discrepancies in findings. The role of short-term versus long-term exposure to air pollution is potentially influencing the results as well. In the present study, we used annual mean concentrations of air pollution as a marker for long-term exposure to air pollution. We did not have access to short-term measurements. It has been proposed that the acute, toxic effects of inflammation following short-term air pollution exposure, such as one’s average daily exposure, may lead to telomere lengthening by the activation of the enzyme telomerase (27). In the present study we find it unlikely that short-term exposure would influence the results however, since we have previously seen that the correlation between short-term levels and long-term exposure is very low in the Betula study (28).

Another potential explanation for the disparity in results in previous studies is the realm of proinflammatory effects of air pollution and differential distribution of leukocytes in response to inflammation. Telomere measurement is conducted as a weighted average of different leukocytes subtypes and it is important to note that proportion of neutrophils in blood circulation is higher than other leukocytes subtypes and they have longer telomere length than lymphocytes (29, 30). Notably, acute inflammation is characterised by predominant accumulataion of neutrophils rather than lymphocytes, thus, it is plausible to presume that inflammation as a result of air pollution exposure may cause longer telomere length due differential clonal expansion of leukocytes (27). In the present study however, we controlled for lymphocyte proportion in an attempt to control for such differential expasions. Anonther mechanism by which air pollution may increase the telomere length is based on inflammation related oxidtative stress. Although, oxidative stress has been suggested to decrease telomere length but contrarily it may also cause an increase in telomere length by pairing up four guanine residues in the DNA strand (31). This enhances the accessibility of telomerase enzyme which eventually causes an increase in telomere length.

There are several strengths and limitations of the present study that should be acknowledged. One of the key strengths is the estimation of air pollution concentrations using a high-resolution dispersion model. These estimates were further refined by accounting for meteorological conditions, traffic volume (including vehicle types and speed), street width and neighbouring building heights. Additionally, this study is among the first, to our knowledge, to examine the effects of source-specific particles on telomere length. Other strengths are the detailed dementia diagnosis and the long follow-up time within the Betula study(15). There are also several limitations to consider. Firstly, telomere length measurements with qPCR are known to suffer from measurement imprecision, although qPCR is still the preferred method in epidemiological studies due to its cost-effectiveness (32). There is furthermore a risk of bias or lack of precision due to self-reported covariate information. There may also be residual confounding. We adjusted for level of education, which previous research has confirmed to increase with telomere length (33), but residual confounding due to socio-economy or lifestyle factors may have biased the estimates. The generalizability of our study is furthermore limited as it focuses on a specific geographic location with relatively low air pollution levels compared to other regions worldwide. The source-specific estimates should be reasonable generalizable to other similar areas in the world, however. Additionally, our study relies on cross-sectional observational data, which makes it challenging to establish causal inferences. However, follow-up for dementia incidence was conducted longitudinally. Moreover, our previous studies in the Betula study shows high correlation between long-term exposure to air pollution and the annual mean (which was used in the present study). We, furthermore, acknowledge the possibility of selection bias, as some participants with short residual leukocyte telomere length (rLTL), which is, again, associated with aging, age-related diseases and mortality, may have died before study enrolment. This could potentially lead to an underestimation of the effect estimates between air pollution and rLTL. Given that the levels of air pollution was low and the study size was limited compared to for example the studies in the UK biobank (22, 23), the statistical power to detect associations could furthermore be too low to detect associations. The particle levels were well above the WHO guidelines for PM2.5 from 2021 however, and in previous studies in Betula we have been able to detect associations between air source-specific pollutants and dementia (21, 34).

## 5. Conclusion

In summary, we did not observe any clear assocition between air pollution and leukocyte telomere length. In people who later developed dementia, there were tendencies for air pollution to be associated with an increase in leukocyte telomere length, however. Despite established health risks associated with air pollution, its specific impact on telomere length is a complex and evolving field. Conflicting study results highlight the need for further investigation to unravel the intricacies and discrepancies, shedding light on underlying mechanisms. Future research should focus on exploring the associations between air pollution exposure and telomere length, with attention to susceptible groups and the mechanisms involved.

## Data Availability

Data are available from the Umeå University for researchers who meet the criteria for access to confidential data. https://www.umu.se/en/research/projects/betula---aging-memory-and-dementia/

## Acknowledgments

The authors are grateful to the Betula study participants.

## Author Contributions

Conceptualization (AO), Formal Analysis (WR), Funding Acquisition (AO, SD, SP, PJ, KMK), Resources (AO, SD, SP, RA), Writing (WR, AO, EF, SP, SD), Writing – Review & Editing (all authors).

## Ethics and Consent

The study was approved by the ethics review authority with Dnr: 2022-04608-01 and written informed consent was given by all Betula participants.

## References

1. Manisalidis I, Stavropoulou E, Stavropoulos A, Bezirtzoglou E. Environmental and health impacts of air pollution: a review. Frontiers in public health. 2020;8:14.

2. WHO. WHO global air quality guidelines: particulate matter (PM2.5 and PM10), ozone, nitrogen dioxide, sulfur dioxide and carbon monoxide. 2021.

3. Abolhasani E, Hachinski V, Ghazaleh N, Azarpazhooh MR, Mokhber N, Martin J. Air pollution and incidence of dementia: A systematic review and meta-analysis. Neurology. 2023;100(2):e242–e54.

4. Livingston G, Huntley J, Sommerlad A, Ames D, Ballard C, Banerjee S, et al. Dementia prevention, intervention, and care: 2020 report of the Lancet Commission. The Lancet. 2020;396(10248):413–46.

5. Scheltens P, De Strooper B, Kivipelto M, Holstege H, Chételat G, Teunissen CE, et al. Alzheimer’s disease. The Lancet. 2021;397(10284):1577–90.

6. Blackburn EH, Epel ES, Lin J. Human telomere biology: a contributory and interactive factor in aging, disease risks, and protection. Science. 2015;350(6265):1193–8.

7. Srinivas N, Rachakonda S, Kumar R. Telomeres and telomere length: a general overview. Cancers. 2020;12(3):558.

8. Hackenhaar FS, Josefsson M, Adolfsson AN, Landfors M, Kauppi K, Hultdin M, et al. Short leukocyte telomeres predict 25-year Alzheimer’s disease incidence in non-APOE ε4-carriers. Alzheimer’s research & therapy. 2021;13(1):1–13.

9. Wang Q, Zhan Y, Pedersen NL, Fang F, Hägg S. Telomere length and all-cause mortality: a meta-analysis. Ageing research reviews. 2018;48:11–20.

10. Zhao B, Vo HQ, Johnston FH, Negishi K. Air pollution and telomere length: a systematic review of 12,058 subjects. Cardiovascular diagnosis and therapy. 2018;8(4):480.

11. Miri M, Nazarzadeh M, Alahabadi A, Ehrampoush MH, Rad A, Lotfi MH, et al. Air pollution and telomere length in adults: A systematic review and meta-analysis of observational studies. Environmental Pollution. 2019;244:636–47.

12. Segersson D, Johansson C, Forsberg B. Near-Source Risk Functions for Particulate Matter Are Critical When Assessing the Health Benefits of Local Abatement Strategies. International journal of environmental research and public health. 2021;18(13):6847.

13. Nilsson L-G, Bäckman L, Erngrund K, Nyberg L, Adolfsson R, Bucht G, et al. The Betula prospective cohort study: Memory, health, and aging. Aging, Neuropsychology, and Cognition. 1997;4(1):1–32.

14. Boraxbekk C-J, Lundquist A, Nordin A, Nyberg L, Nilsson L-G, Adolfsson R. Free Recall Episodic Memory Performance Predicts Dementia Ten Years prior to Clinical Diagnosis: Findings from the Betula Longitudinal Study. Dementia and Geriatric Cognitive Disorders Extra. 2015;5(2):191–202.

15. Nyberg L, Boraxbekk C-J, Sörman DE, Hansson P, Herlitz A, Kauppi K, et al. Biological and environmental predictors of heterogeneity in neurocognitive ageing: evidence from Betula and other longitudinal studies. Ageing Research Reviews. 2020:101184.

16. Cawthon RM. Telomere measurement by quantitative PCR. Nucleic acids research. 2002;30(10):e47-e.

17. Pudas S, Josefsson M, Nordin Adolfsson A, Landfors M, Kauppi K, Veng-Taasti LM, et al. Short leukocyte telomeres, but not telomere attrition rates, predict memory decline in the 20-year longitudinal betula study. The Journals of Gerontology: Series A. 2021;76(6):955–63.

18. Segersson D, Eneroth K, Gidhagen L, Johansson C, Omstedt G, Nylén AE, et al. Health impact of PM10, PM2. 5 and black carbon exposure due to different source sectors in Stockholm, Gothenburg and Umea, Sweden. International journal of environmental research and public health. 2017;14(7):742.

19. Hausberger S. Emission Factors from the Model PHEM for the HBEFA Version 3. 2009.

20. Omstedt G, Forsberg B, Persson K. Wood Smoke in Västerbotten—Mesurements, Calculations and Health Impact. SMHI; 2014.

21. Andersson J, Sundström A, Nordin M, Segersson D, Forsberg B, Adolfsson R, et al. PM 2.5 and Dementia in a Low Exposure Setting: The Influence of Odor Identification Ability and APOE. Journal of Alzheimer’s Disease. 2023(Preprint):1–11.

22. Wu Y, Gasevic D, Wen B, Yu P, Xu R, Zhou G, et al. Association between air pollution and telomere length:: A study of 471,808 UK Biobank participants. The Innovation Medicine. 2023;1(2):100017.

23. Bountziouka V, Hansell AL, Nelson CP, Codd V, Samani NJ. Large-Scale Analysis of the Association between Air Pollutants and Leucocyte Telomere Length in the UK Biobank. Environmental Health Perspectives. 2023;131(2):027701.

24. Walton RT, Mudway IS, Dundas I, Marlin N, Koh LC, Aitlhadj L, et al. Air pollution, ethnicity and telomere length in east London schoolchildren: an observational study. Environment international. 2016;96:41–7.

25. Xia Y, Chen R, Wang C, Cai J, Wang L, Zhao Z, et al. Ambient air pollution, blood mitochondrial DNA copy number and telomere length in a panel of diabetes patients. Inhalation toxicology. 2015;27(10):481–7.

26. Wang C, Wolters PJ, Calfee CS, Liu S, Balmes JR, Zhao Z, et al. Long-term ozone exposure is positively associated with telomere length in critically ill patients. Environment international. 2020;141:105780.

27. Hou L, Wang S, Dou C, Zhang X, Yu Y, Zheng Y, et al. Air pollution exposure and telomere length in highly exposed subjects in Beijing, China: a repeated-measure study. Environment international. 2012;48:71–7.

28. Andersson J, Oudin A, Nordin S, Forsberg B, Nordin M. PM2. 5 exposure and olfactory functions. International Journal of Environmental Health Research. 2022;32(11):2484–95.

29. Fernandes SG, Dsouza R, Khattar E. External environmental agents influence telomere length and telomerase activity by modulating internal cellular processes: Implications in human aging. Environmental Toxicology and Pharmacology. 2021;85:103633.

30. Aubert G, Baerlocher GM, Vulto I, Poon SS, Lansdorp PM. Collapse of telomere homeostasis in hematopoietic cells caused by heterozygous mutations in telomerase genes. PLoS genetics. 2012;8(5):e1002696.

31. Lee H-T, Bose A, Lee C-Y, Opresko PL, Myong S. Molecular mechanisms by which oxidative DNA damage promotes telomerase activity. Nucleic acids research. 2017;45(20):11752–65.

32. Verhulst S, Susser E, Factor-Litvak PR, Simons MJ, Benetos A, Steenstrup T, et al. Commentary: The reliability of telomere length measurements. International journal of epidemiology. 2015;44(5):1683–6.

33. Amin V, Fletcher JM, Sun Z, Lu Q. Higher educational attainment is associated with longer telomeres in midlife: Evidence from sibling comparisons in the UK Biobank. SSM-Population Health. 2022;17:101018.

34. Oudin A, Segersson D, Adolfsson R, Forsberg B. Association between air pollution from residential wood burning and dementia incidence in a longitudinal study in Northern Sweden. PloS one. 2018;13(6):e0198283.

